# IRAK2 deficiency causes a new immune dysregulation disorder

**DOI:** 10.1101/2024.10.21.24315796

**Authors:** Yudie Fei, Lin Liu, Shuangyue Ma, Shihao Wang, Meiping Lu, Jing Xue, Ying Jin, Yusha Wang, Xiangwei Sun, Xiang Chen, Xu Han, Changming Zhang, Li Guo, Jiahui Zhang, Hua Zhong, Lihong Wen, Xiaomin Yu, Qing Zhou, Zhihong Liu

**Affiliations:** Liangzhu Laboratory, Zhejiang University, Hangzhou, China; Urology & Nephrology Center, Department of Nephrology, Zhejiang Provincial People’s Hospital (Affiliated People’s Hospital, Hangzhou Medical College), Hangzhou, China; Department of Rheumatology Immunology and Allergy, Children’s Hospital, Zhejiang University School of Medicine, Hangzhou, China; Department of Rheumatology, The Second Affiliated Hospital, Zhejiang University School of Medicine, Hangzhou, Zhejiang, China; National Clinical Research Center for Kidney Diseases, Jinling Hospital, Affiliated Hospital of Medical School, Nanjing University, Nanjing, China; Cell and Molecular Biology Laboratory, Affiliated Zhoushan Hospital of Wenzhou Medical University, Zhoushan, China

**Author notes:** Yudie Fei, Lin Liu, Shuangyue Ma, Shihao Wang, Meiping Lu, Jing Xue, Ying Jin contributed equally to this work.

## Abstract

Interleukin 1 receptor-associated kinase 2 (IRAK2) plays a critical role in immune response by participating in the formation of the Myddosome complex in Toll-like receptor (TLR) signaling pathways. Here, we identified a loss-of-function mutation (*IRAK2-Δex2*) in the *IRAK2* gene in three patients, presenting with immune dysregulation, including systemic lupus erythematosus (SLE) and autoinflammatory disease. This mutation leads to the skipping of exon 2 in *IRAK2*, disrupting its interaction with IRAK4 and impairing the activation of nuclear factor kappa B (NF-*κ*B) and mitogen-activated protein kinase (MAPK) signaling pathways via Myddosome. The patients exhibited aberrantly upregulated type I interferon (IFN) response following LPS stimulation, which was further confirmed in bone marrow-derived macrophages (BMDMs) in mice. Our study suggests that IRAK2 deficiency results in immune dysregulation due to compromised TLR signaling and activated IFN signaling primarily in monocyte-macrophage lineage.

**One Sentence Summary:** A new immune dysregulation disorder caused by a loss-of-function mutation in the *IRAK2* gene, which disrupts TLR signaling via Myddosome, results in impaired NF-*κ*B activation and upregulated type I interferon responses.

## INTRODUCTION

TLRs are classic pattern recognition receptors (PRR) that play an indispensable role in immune defense, as well as mediating cross-talk between the innate and adaptive immune systems. TLRs are activated by pathogen-associated molecular patterns (PAMPs) or damage-associated molecular patterns (DAMPs) (*1, 2*). Upon ligand recognition, myeloid differentiation factor 88 (MyD88), interleukin 1 receptor-associated kinase 4 (IRAK4), and IRAK2 are recruited to the membrane to form the Myddosome complex (*3*), which primarily activates the NF-*κ*B and MAPK signaling pathways (*4*). Notably, TLR4, the first TLR identified in mammals (*5, 6*), recognizes lipopolysaccharide (LPS) and activates not only NF-*κ*B and MAPK pathways but also type I IFN signaling (*7, 8*).

IRAK2, composed of a death domain (DD), a proline, serine, and threonine-rich (ProST) linker, a kinase domain (KD), and a C-terminal domain (CD), is a key component of the Myddosome (*3, 9*), which is involved in most TLR signaling pathways except TLR3 (*10*). Furthermore, IRAK2 shares functional redundancy with IRAK1. Previous studies have shown that IRAK1 and IRAK2 sequentially regulate TLR signaling (*11*). While single knockout of either *Irak1* or *Irak2* only partially impairs TLR signaling, double knockouts of both *Irak1* and *Irak2* completely silence TLR signaling, resembling the effects of *Irak4* knockout in mice (*11*). In humans, *IRAK2* has only one transcript, whereas in mice, there are four transcripts, two of which lack exon 2 (*12*). Exon 2 encodes a proportion of the DD, a critical protein domain for Myddosome assembly (*3*). Transcripts that lack exon 2 exhibit an inhibitory effect on signal transduction. Several pathogenic genes have been identified in the TLR signaling pathway. Patients with loss-of-function mutations in either *MyD88* or *IRAK4* are highly susceptible to pyogenic infections due to impaired TLR and interleukin 1 (IL-1) signaling (*13, 14*). Additionally, many patients with IRAK4 deficiency also develop autoimmune disease (*15–17*), suggesting a more complex pathological mechanism of Myddosome deficiency. However, IRAK2 deficiency has not been reported in humans. Here, our study is the first report of a loss-of-function mutation in *IRAK2* causing immune dysregulation.

## RESULTS

### *IRAK2-Δex2* mutation in patients with immune dysregulation disorders

The proband (P1) of the first family (Fig. 1A) is a female in her 40s who first presented with swelling of both lower limbs, facial erythema, arthralgia, proteinuria, and thrombocytopenia in her 20s. Prednisone and cyclophosphamide (CTX) were used and her proteinuria gradually returned negative. She has experienced recurrent episodes of disease, which presented with proteinuria, hematuria, pancytopenia, positive autoantibodies and decreased C3 (table S1). Based on these findings, she was diagnosed with SLE, following the European Alliance of Associations for Rheumatology/American College of Rheumatology classification criteria (*18*). A kidney biopsy revealed class IV+V lupus nephritis (LN) (Fig. 1B and fig. S1A). Prednisone combined with CTX pulse treatment (9.3 g in total) resulted in partial remission. Then prednisone and azathioprine (AZA) or mycophenolate mofetil (MMF) were prescribed. Complete remission was achieved and maintained until the last visit. Contrary to her autoimmune symptoms, she was diagnosed with hypogammaglobulinemia (table S1). During the pandemic, she experienced a severe COVID-19 infection. P2 has not yet developed symptoms but occasionally experiences skin rashes (fig. S1A). Her laboratory tests revealed positive ANA levels, which has been increasing over time (table S2), along with other autoantibodies, including Anti-β2 glycoprotein (A-β2-GP1) and anticardiolipin antibodies (ACA). Both P1 and P2 exhibited an increased CD8+ T cells population (table S3).

**Fig. 1.**
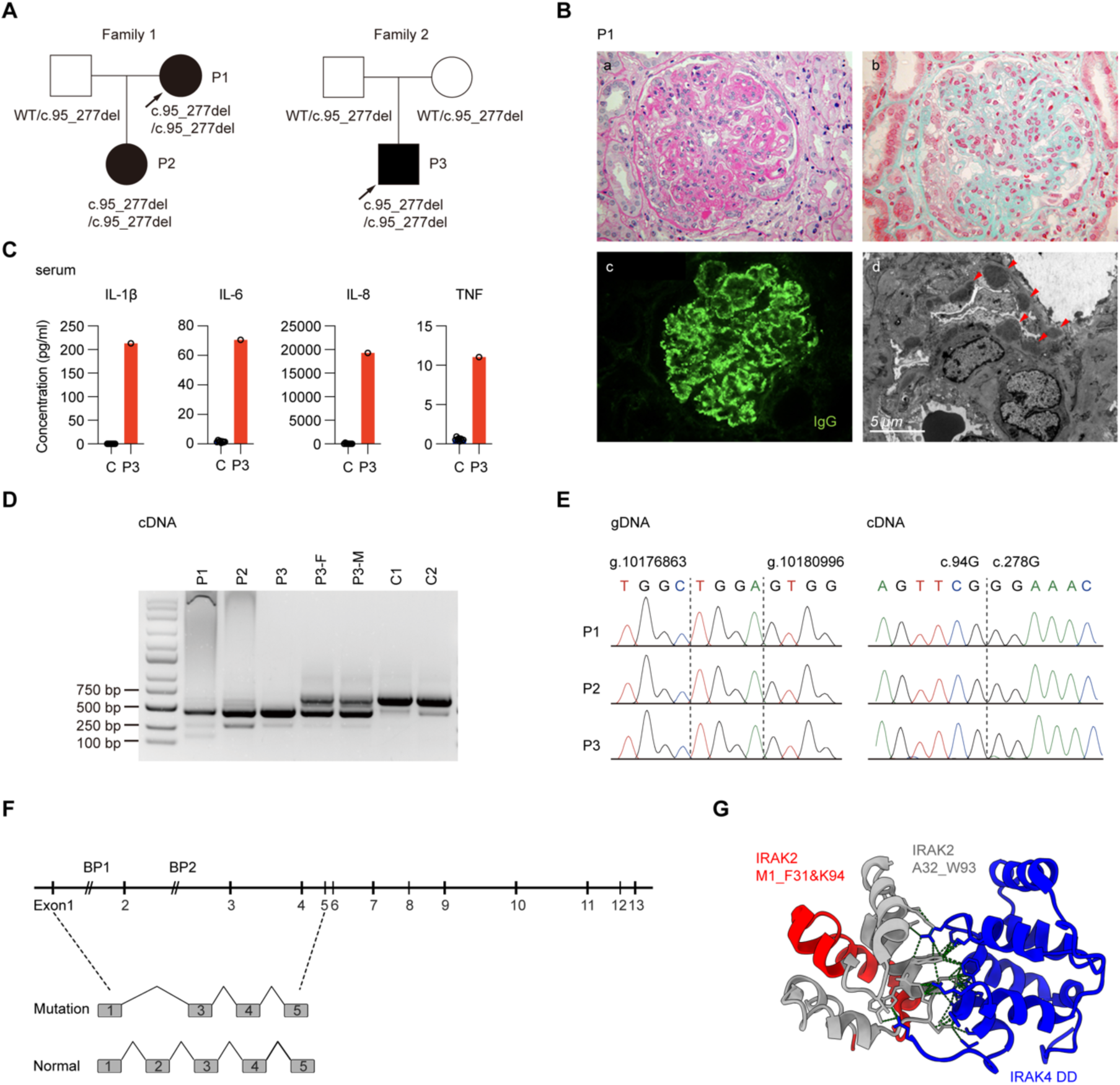
Clinical manifestations of the patients and identification of *IRAK2-*Δ*ex2* mutation in two unrelated families. (A) Pedigrees of the two unrelated families carrying the *IRAK2-Δex2* mutation. The probands are indicated by an arrow. (B) Renal pathological examination of P1. Light microscopy: (a) A cellular crescent compressing the tuft of the glomerulus, with diffuse mesangial and endocapillary proliferation in the glomerulus (PAS, ×400). (b) Eosinophilic deposits are observed in the mesangial area and subendothelial segments, as well as on the epithelial side. These deposits show segmental peripheral stratification and the formation of segmental “spikes” (Masson, ×400). Immunofluorescence: (c) IgG deposits diffusely distributed in a granular pattern in the vascular loops (IF, ×400). Electron microscopy: (d) Abundant electron-dense deposits (indicated by arrowheads) in the mesangial area. (C) Levels of proinflammatory cytokines IL-1β, IL-6, TNF, and chemokine IL-8 in the serum of P3 and eight unaffected controls detected by CBA. Bars represent the mean ± SD. (D) Agarose gel electrophoresis results of PCR amplification products targeting Exon 2 of *IRAK2* using cDNA as templates. Confirmation of the variant by Sanger sequencing. (F) Schematic diagram of the transcription pattern in the presence or absence of the variant. (G) Structure of DD of IRAK4 (blue) and IRAK2 (gray indicates the missing part and red indicates the remaining part) (PDB 3MOP). Green dotted lines indicate the intermolecular interactions at the contact interface of the two molecules. Kidney biopsy was performed on P1 during her hospitalization. PAS, periodic acid-Schiff.

P3 is a male in his 20s who has experienced recurrent episodes of fever, oral ulcers, genital ulcers, skin lesions, and proteinuria. Additionally, gastrointestinal endoscopy revealed multiple ulcers in the terminal ileum of P3. He was diagnosed with autoinflammatory disease. His laboratory tests showed elevated C-reactive protein (CRP) and erythrocyte sedimentation rate (ESR) during febrile episodes. This was further supported by the detection of proinflammatory cytokines using Cytometric Bead Array (CBA) (Fig. 1C).

Whole exome sequencing (WES) of P1and P3 identified a homozygous deletion of exon 2 in the *IRAK2* gene (c.95_277del, p.Ala32_Trp93delinsGly) (fig. S1B), which was confirmed by Sanger sequencing, revealing that both families share the same breakpoints (fig. S1C). RT-PCR and Sanger sequencing confirmed this as an in-frame deletion (Fig. 1, D, E, and F), resulting in a partial deletion of the DD of the IRAK2 protein. All three patients are homozygous for the *IRAK2-Δex2* mutation, while the other family members in the trio are heterozygous for this mutation. No other rare variants of unknown significance or copy number variants (CNVs) in genes previously associated with immune disorders were found in either family. Additionally, this variant has not been reported in any public database of human exomes or Chinese cohorts. Previous studies have elucidated the structure of Myddosome (*3*). Visualization using Chimera X (*19*) reveals that the skipping region of DD in IRAK2 predominantly resides on the interaction surface with IRAK4 (Fig. 1G). The *IRAK2-Δex2* mutation is similar to the inhibitory transcripts found in mice, which also lack exon 2 (*12*).

### Disrupted interaction between the IRAK2-Δex2 mutation and IRAK4

The direct interaction between IRAK2 and IRAK4 is critical for signal transduction of all TLRs except TLR3 (*10*). Given that the *IRAK2-Δex2* mutation leads to a partial deletion of the DD in IRAK2 protein, we conducted co-immunoprecipitation experiments by overexpressing IRAK2 along with either IRAK4 or tumor necrosis factor receptor-associated factor 6 (TRAF6) in HEK293T cells. The results showed that the mutation disrupted the interaction between IRAK2 and IRAK4, but with no significant change in its interaction with TRAF6 (Fig. 2A), which were consistent with the predicted impact of the mutation site.

**Fig. 2.**
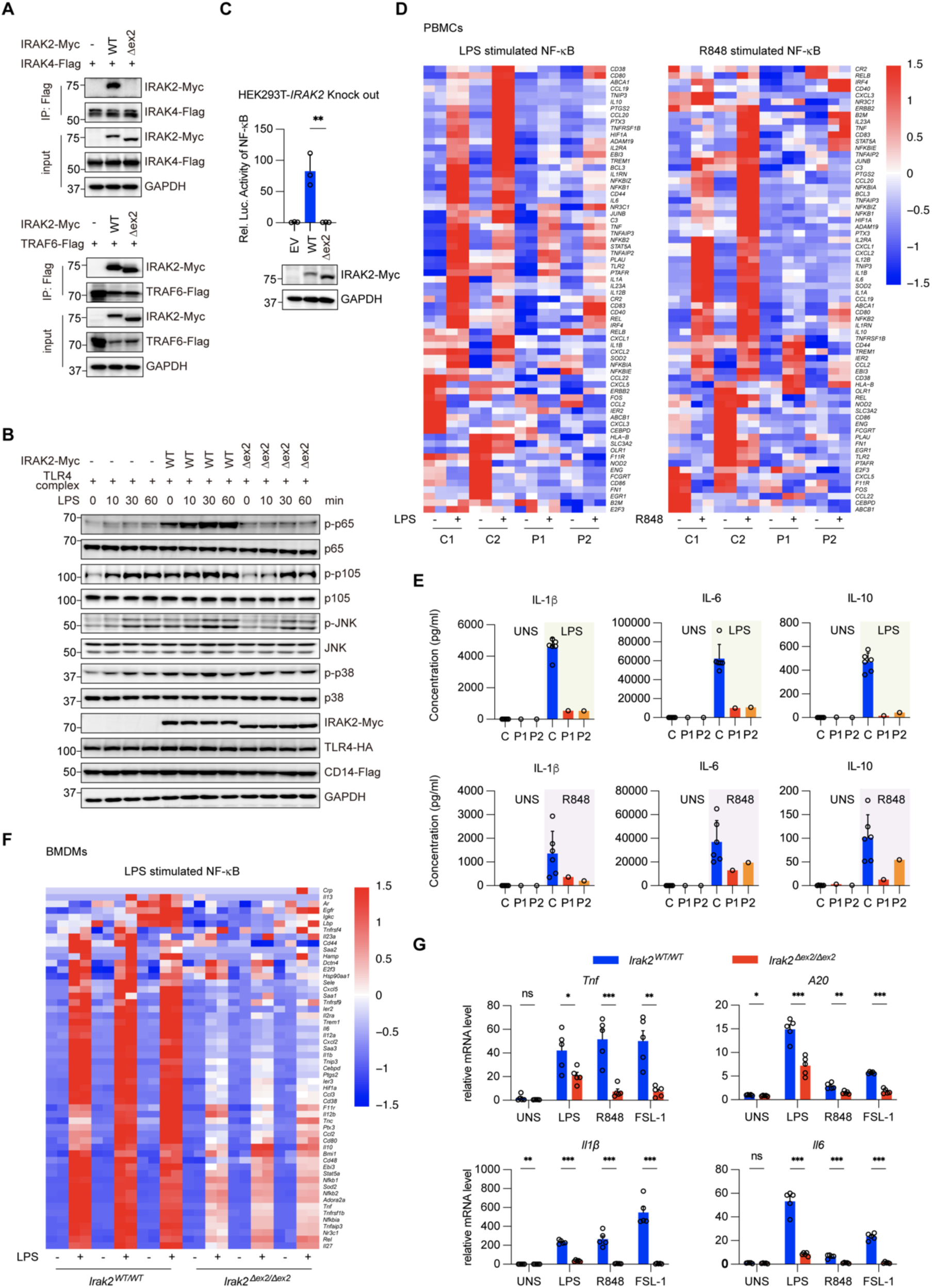
Impaired interaction between IRAK4 and the IRAK2-Δex2 mutation results in deficient signaling pathways via Myddosome. (A) Co-immunoprecipitation of whole cell lysates from HEK293T cells transiently expressing Myc-tagged IRAK2-WT or IRAK2-Δex2 and equal levels of IRAK4-Flag or TRAF6-Flag using anti-Flag antibody (IP: Flag). The experiment was repeated three times. (B) Immunoblot showing deficiency of NF-*κ*B and MAPK signaling in the presence of IRAK2-Δex2 mutation compared with IRAK2-WT, using indicated antibodies in the stable HEK293T-TLR4 cell line. The experiment was repeated three times. (C) NF-*κ*B signaling was detected by luciferase assay in HEK293T-*IRAK2^−/−^* cells overexpressing either IRAK2-WT or IRAK2-Δex2. Cells were co-transfected with NF-*κ*B-luciferase reporter and renilla plasmids (n=3). Bars represent the mean ± SD. An unpaired Student’s *t*-test was performed between the IRAK2-WT and IRAK2-Δex2 groups, \*\**P* < 0.01. The experiment was repeated three times. (D) RNA sequencing analysis of NF-*κ*B pathway in PBMCs treated with 1 μg/ml LPS, 1 μg/ml R848, respectively, or left untreated for 12 hours. (E) PBMCs from P1, P2, and six unaffected controls were treated with 1 μg/ml LPS, 1 μg/ml R848, respectively, or left untreated for 12 hours. Supernatants were collected for CBA. Bars represent the mean ± SD. (F) RNA sequencing analysis of NF-*κ*B pathway in BMDMs treated with 100 ng/ml LPS or left untreated for 8 hours (n=3). (G) Quantitative PCR (qPCR) analysis of the expression of the genes related to NF-*κ*B signaling in BMDMs treated with 100 ng/ml LPS, 25 ng/ml R848, or 50 ng/ml FSL-1, respectively, or left untreated for 8 hours (n=5). Bars represent the mean ± SEM. Unpaired Student’s *t*-tests were performed between the *Irak2^WT/WT^* and *Irak2*^Δ^*^ex2/^*^Δ^*^ex2^* groups, \**P* < 0.05, \*\**P* < 0.01, \*\*\**P* < 0.001. UNS, unstimulated; ns, not significant.

### IRAK2-Δex2 mutation disrupts signal transduction via Myddosome

To study the functional changes in signal transduction, we overexpressed either wild-type IRAK2 (IRAK2-WT) or IRAK2-Δex2 in a stable HEK293T-TLR4 cell line, which continuously expresses TLR4-HA, CD14-Flag, and LY96 (*20*). After stimulating the TLR4 complex, IRAK2-Δex2 failed to enhance the phosphorylation of p65, p105, JNK, and p38 compared with IRAK2-WT (Fig. 2B). We then generated HEK293T-*IRAK2^−/−^* cells using CRISPR-Cas9 techniques (*21*) to exclude the interference from endogenously expressed IRAK2, and *IRAK2^−/−^* cells were unable to activate NF-*κ*B signaling pathway, as confirmed by immunoblot and qPCR (fig. S2, A and B). Additionally, luciferase assay results showed that IRAK2-Δex2 failed to activate the NF-*κ*B signaling pathway compared with IRAK2-WT (Fig. 2C).

To investigate the functional impact of the IRAK2-Δex2 mutation in patients, PBMCs from patients and unaffected controls were stimulated with LPS or TLR7/8 agonist R848. RNA sequencing analysis indicated that PBMCs from the two patients consistently exhibited defects in activating NF-*κ*B signaling in response to LPS or R848 stimulation (Fig. 2D and fig. S2C), as well as impaired activation of the MAPK signaling pathway (fig. S2D). Furthermore, proinflammatory cytokines downstream of the NF-*κ*B pathway, including IL-1β, IL-6, and IL-10, did not show induction following LPS or R848 stimulation in patients’ PBMCs (Fig. 2E).

Moreover, homozygous knock-in *Irak2^Δex2/Δex2^*mice were generated on a C57BL/6 background to further explore the physiological function of the IRAK2-Δ ex2 mutation. RNA sequencing demonstrated that BMDMs from *Irak2*^Δ^*^ex2/^*^Δ^*^ex2^* mice exhibited defects in NF-*κ*B and MAPK signaling pathways, similar to patients’ PBMCs (Fig. 2F and fig. S2E). Moreover, qPCR analysis revealed that in addition to defects in the Tlr4 and Tlr7/8 pathways, the Tlr2/Tlr6 pathway was also impaired following FSL-1 stimulation (Fig. 2G). These findings suggest that IRAK2-Δex2 mutation causes a broad deficiency in signaling pathways via Myddosome.

### Impaired NF-κB signaling in patients’ myeloid cells

To systematically analyze the inflammatory response at the single-cell level, we performed Cytometry by time of flight (CyTOF) on PBMCs from patients and unaffected controls. PBMCs were treated with LPS or untreated for 12 hours prior to CyTOF analysis. No significant changes were observed in the composition of PBMCs with or without LPS stimulation (Fig. 3, A and B and fig. S3A). CyTOF demonstrated a decreased induction of proinflammatory cytokines, including IL-6 and IL-10, in monocytes and dendritic cells (DCs) from patients compared with unaffected controls following LPS stimulation (Fig. 3C).

**Fig. 3.**
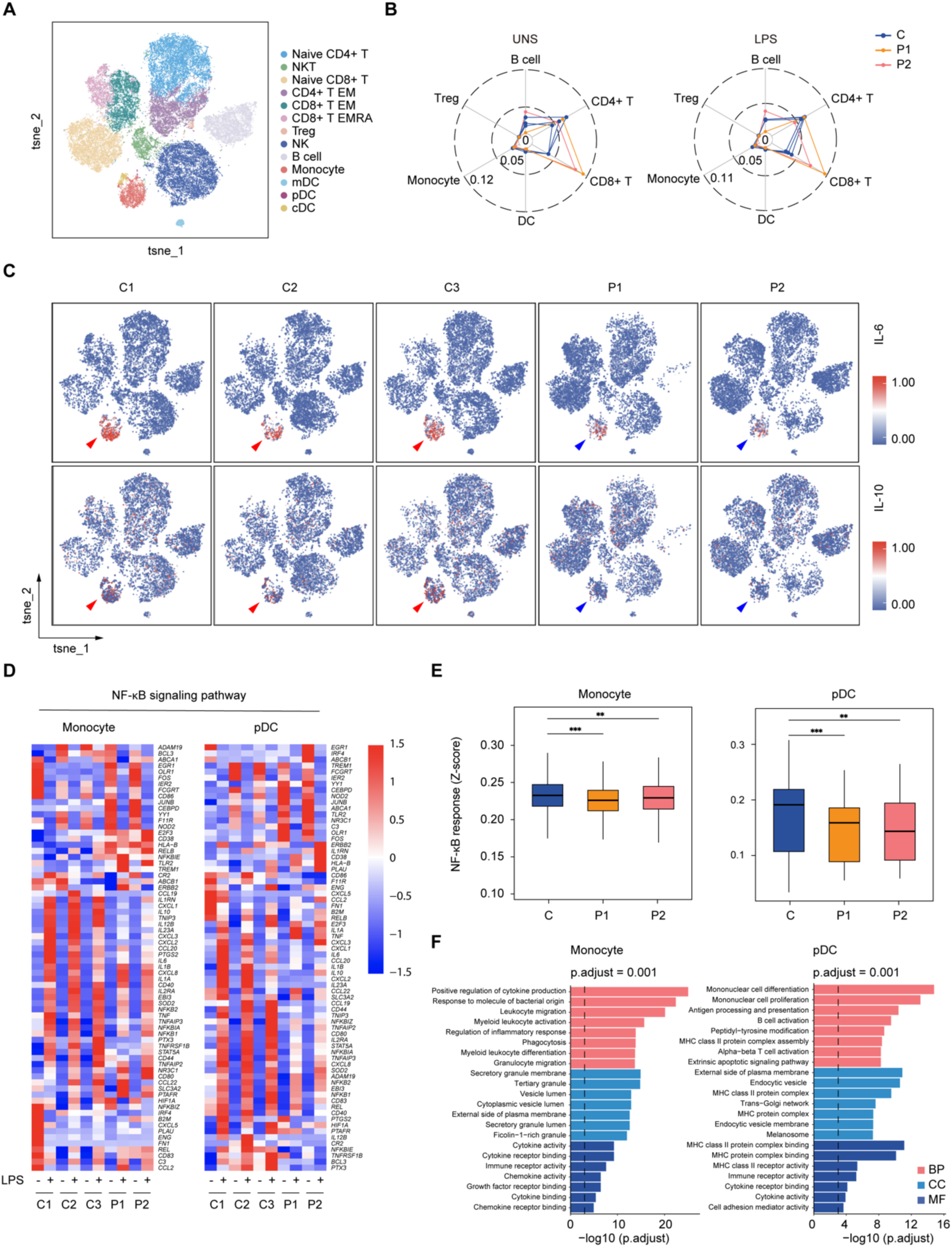
Deficiency in NF-*κ*B signaling in myeloid cells in patients’ PBMCs following LPS stimulation. (A) The t-distributed stochastic neighbor embedding (t-SNE) representation illustrates the immunophenotyping of 5*10,000 CD45^+^ CD66b^−^ live PBMCs treated with 100 ng/ml LPS for 12 hours from two patients and three unaffected controls, as analyzed by CyTOF. (B) The distribution of immune cells for two patients and three unaffected controls under untreated conditions (left) and LPS stimulating condition (right). (C) Expressions of IL-6 and IL-10 are displayed on a tSNE map of 10,000 live PBMCs from the indicated individuals. Arrow heads denotes the clusters of monocytes, conventional dendritic cells (cDC), and plasmacytoid dendritic cells (pDC). (D) Comparison of NF-*κ*B signaling pathway in monocytes and pDC subsets from P1, P2, and three unaffected controls, as determined by scRNA-seq. (E) The box plots show the NF-*κ*B response scores calculated by scRNA-seq for monocytes and pDC subsets. Statistical analysis was performed using Wilcoxon test (\*\**P* < 0.01, \*\*\**P* < 0.001). (F) Functional enrichment of monocytes and pDC is performed using scRNA-seq data, gene sets are obtained from Gene Ontology categories, including Biological Process (BP), Cellular Component (CC), and Molecular Function (MF). Enrichment results were filtered by adjusted p-value (method = “BH”).

Subsequently, single-cell RNA sequencing (scRNA-seq) was performed to estimate cell-type specific immune signatures. Consistent with CyTOF results, impaired NF-*κ*B signaling was observed in monocytes and pDCs after LPS stimulation (Fig. 3, D and E). This highlights the critical role of monocytes and pDCs in the innate immune system. Functional enrichment analysis (Geno Ontology gene sets) indicated abnormal activation of monocytes and pDCs in the patients (Fig. 3F), suggesting aberrantly activated inflammatory response in the presence of impaired NF-*κ*B signaling. Collectively, these findings indicate that the myeloid cells from patients failed to respond to LPS stimulation, and suggest involvement of additional signaling pathways alongside impaired NF-*κ*B signaling.

### Elevated type I IFN response following LPS stimulation

We performed gene set enrichment analysis (GSEA) (*22*) on bulk RNA sequencing data of PBMCs to explore aberrantly activated signaling pathways associated with the IRAK2-Δex2 mutation. The analysis revealed upregulation of interferon α and interferon γ responses (Fig. 4A), suggesting activation of interferon signaling pathway in patients. Quantification of 28-type I interferon signature score (*23*) from the bulk RNA sequencing data demonstrated that patients had significantly higher scores compared with unaffected controls under both untreated and LPS-treated conditions (Fig. 4B), confirming the increased interferon signatures in patients with IRAK2 deficiency.

**Fig. 4.**
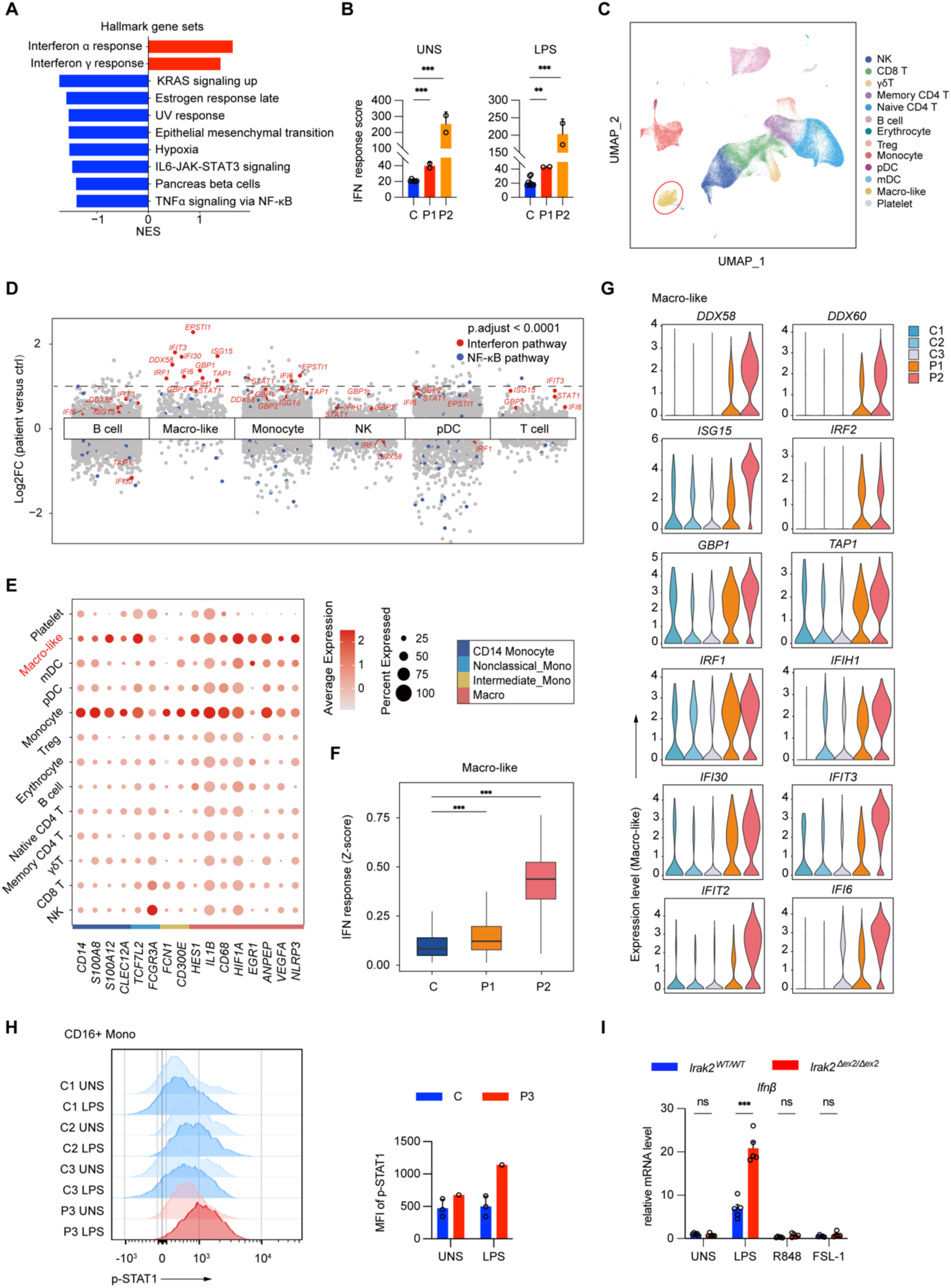
Elevated type I IFN signature following LPS stimulation in the presence of IRAK2-Δ ex2 mutation. (A) The gene set enrichment analysis (GSEA) of RNA sequencing data for PBMCs. NES, normalized enrichment score. (B) Quantification of the 28-gene IFN score of RNA sequencing data of total RNA extracted from PBMCs. Data are presented as the mean ± SD. Significance assessed by unpaired Student’s *t*-test, \*\*\**P* < 0.001, \*\**P* < 0.01. (C) Uniform Manifold Approximation and Projection (UMAP) visualization and marker-based annotation of 13 cell subtypes from P1, P2, and three unaffected controls. ‘Macro-like’ cell subset is circled in red. NK, natural killer cells; Treg, regulatory T cells; pDC, plasmacytoid dendritic cells; mDC, myeloid dendritic cells; Macro-like: macrophage-like. (D) Identification of differentially expressed genes between patients (P1 and P2) and three unaffected controls across different cell types. Each point denotes a gene with an adjusted p-value (method= “BH”) less than 0.05. The dashed line represents a Log2FC value of 1, marking the number of differentially expressed genes exceeding this threshold across cell clusters. (E) The bubble graph shows the expression of marker genes for annotating monocytes and ‘Macro-like’ cell subset by scRNA-seq. The size of the bubble indicates the proportion of cells expressing the gene, and the color scale indicates the average expression level. (F) The box plot represents the type I interferon response scores calculated by scRNA-seq for ‘Macro-like’ cell subset. Statistical analysis was performed using Wilcoxon test (\*\*\**P* < 0.001). (G) The violin plots show the upregulation of IFN-stimulated genes in the ‘Macro-like’ cell subset of P1 and P2 compared to three unaffected controls. (H) The phosphorylation level of STAT1 of CD16+ monocytes within PBMCs from P3 and three unaffected controls, as determined by flow cytometry analysis. (I) qPCR analysis of *Ifnβ* expression in BMDMs treated with 100 ng/ml LPS, 25 ng/ml R848, or 50 ng/ml FSL-1, respectively, or left untreated for 8 hours (n=5). Bars represent the mean ± SEM. An unpaired Student’s *t*-test was conducted between *Irak2^WT/WT^* and *Irak2*^Δ^*^ex2/^*^Δ^*^ex2^* groups, \*\*\**P* < 0.001.

We performed cell clustering based on scRNA sequencing data and observed a reduced proportion of monocytes following LPS stimulation (fig. S3B). Additionally, a specific cell group was identified in both patients (P1 and P2) and unaffected controls (Fig. 4C and fig. S3B). Genes involved in interferon signaling were significantly upregulated in this specific cell group in patients (Fig.4D). We then examined the cell markers used for annotating clusters. The results indicated that marker genes associated with the myeloid cell lineage, specifically for monocytes and macrophages (*24*)), including *CD68*, *HIF1A*, *EGR1*, *ANPEP*, *VEGFA*, and *NLRP3*, were highly expressed in this specific cell group (Fig. 4E, fig. S3D). Furthermore, we employed the CytoTRACE algorithm (*25*) to assess the differentiation state of monocytes and this specific cell group. This cell group exhibited lower predicted scores than monocytes, indicating a more differentiated state (fig. S3C). Consequently, we defined this cell group as ‘Macrophage-like’ (‘Marco-like’) cell subset, and elevated interferon signaling in patients’ ‘Macro-like’ cell subset was confirmed through AUCell function in the scRNA-seq pipeline (Fig. 4F and fig. S3, E and F) (*26*). Moreover, specific genes in the type I IFN pathway were significantly upregulated in the ‘Macro-like’ cell subset following LPS stimulation (Fig. 4G). Macrophages play important roles in sensing stimuli and regulating immune responses (*27, 28*). These findings suggest that the ‘Macro-like’ cell subset may play a role in exacerbating inflammation through type I IFN pathway. To confirm these findings, flow cytometry revealed stronger phosphorylation of STAT1 in P3’s CD16+ monocytes compared with unaffected controls (Fig. 4H). Additionally, transcription of *Ifnβ* was significantly upregulated in *Irak2^Δex2/Δex2^*mice after LPS stimulation (Fig. 4I). Together, these findings suggest that the IRAK2-Δex2 mutation results in upregulated interferon signaling, primarily in monocyte-macrophage lineage.

## DISCUSSION

Our study identifies a novel homozygous loss-of-function mutation, *IRAK2-Δex2*, associated with immune dysregulation disorders. P1 was diagnosed with SLE, while P3 presented with autoinflammatory disease. Despite the varying clinical manifestations, all three patients exhibited a diminished NF-*κ*B signaling and an upregulated type I IFN signature. Such clinical heterogeneity is also observed in other autoinflammatory diseases. For instance, patients with haploinsufficiency A20 (HA20) display early-onset autoimmune or autoinflammatory diseases, including SLE and Behçet’s disease (*29, 30*).

*IRAK2-Δex2* mutation is a CNV alteration, which encompasses the entire exon 2 and portions of the flanking introns. Notably, this region is known as an *Alu*-mediated homologous recombination hotspot (*31, 32*), which likely contributes to the mechanistic basis of this mutation. Due to variable length, repetitive sequences, and the limitation in sequencing technologies, the detection rate of CNVs is lower than that of single nucleotide polymorphisms (SNPs) (*33, 34*), which hindered the exploration of genetic diseases and the implementation of precision medicine. As CNVs are increasingly recognized as contributors to genetic diseases (*35*), focusing on specific genomic regions, such as homologous recombination hotspots, for screening rare pathogenic variants may prove to be an effective strategy. Although the disease is caused by a homozygous mutation, Family 1 exhibits pseudo-dominant inheritance of the mutation. Family 1 originates from a small town in China, where the founder effect may have contributed to a higher mutation frequency in this population. This observation highlights the need to expand genetic screening in Chinese population, with particular emphasis on the genomic region surrounding *IRAK2*.

The IRAK2-Δex2 mutation results in a large deletion of DD, which abolishes its interaction with IRAK4. Consequently, the assembly of the Myddosome is disrupted. Therefore, cells expressing IRAK2-Δex2 exhibit a broad deficiency in TLR signaling pathways through Myddosome compared with those expressing IRAK2-WT, further validating the mutation as a loss-of-function mutation. CyTOF and scRNA-seq reveal that myeloid cells, such as monocytes and DCs, exhibit impaired NF-*κ*B signaling in patients. Impaired TLR signaling has been associated with autoimmunity and autoinflammation. Previous study has reported that patients with biallelic *TLR4* deficiency exhibit an inborn error of immunity in responding to LPS, with some patients developing Crohn’s disease (*36*). And patients with IRAK4 deficiency, initially diagnosed with primary immunodeficiency (PID) (*14*), subsequently develop autoimmune and autoinflammatory diseases (*15–17*). These observations suggest that impaired TLR signaling via Myddosome contributes to the development of autoimmunity or autoinflammation. In our study, all patients exhibited signs of autoimmunity or autoinflammation. NF-*κ*B signaling, a key pathway downstream of TLRs, is a critical mediator of the inflammatory response and plays paradoxical roles in the regulation of autoimmunity and inflammation. Proper NF-*κ*B signaling is essential for the maintenance of immune tolerance (*37*). These findings highlight the complex interplay between NF-*κ*B signaling, immune homeostasis, and inflammation. Further research is urgently needed to elucidate the intricate signaling networks and immune cell developmental processes involved in these diseases.

Despite reduced NF-*κ*B signaling, an aberrantly upregulated IFN response was observed in patients’ PBMCs and BMDMs of *Irak2^Δex2/Δex2^* mice following LPS stimulation. Notably, P1 is the oldest patients in our study and has developed SLE. Importantly, SLE is significantly associated with type I IFN signaling, with disease activity and severity positively correlating with type I interferon levels (*38*). Type I interferonopathy was first recognized in 2011 (*39*) and has generated increasing attention over the past decade. Advances in high-throughput sequencing technology have facilitated the identification of numerous pathological genes associated with type I interferonopathy. Disturbances in either DNA-mediated signaling (*TREX1*, the RNase H2 complex, *SAMHD1*, and the U7 small nuclear RNP complex) or RNA-mediated signaling (*ADAR1*, *MDA5*) can lead to Aicardi-Goutières syndrome (AGS), which shares clinical manifestations with SLE (*40–44*). Gain-of-function mutations in *TMEM173* result in STING-associated vasculopathy with onset in infancy (SAVI), characterized by early systemic inflammation, vasculopathy, and a range of complications, including severe skin rashes, lung disease, and neurological issues (*45*). Moreover, the *DDX58 R109C* mutation, found in patients with lupus nephritis, leads to hyperactivation of RIG-I and upregulation of type I IFN signaling (*46*). Our experiments and analysis also decipher the correlation of diseases and abnormal activation of type I IFN pathway. ScRNA-seq revealed that the IFN signature was primarily upregulated in a subset of cells resembling macrophages, which underwent transcriptional changes. This underscores the critical role of the monocyte-macrophage lineage in the pathology of the disease.

Although we have confirmed the pathogenicity of the *IRAK2-Δex2* mutation and identified an abnormally activated type I IFN signature in both patients and mice, the direct trigger of this dysregulated signaling remains unclear. Furthermore, the relationship between diminished NF-*κ*B signaling via Myddosome and the elevated type I IFN signature is not yet fully understood. These challenges are commonly encountered in cases of immune dysregulation associated with NF-*κ*B deficiency.

Importantly, genetic screening of IRAK2 deficiency provides potential benefits for prognosis by facilitating early diagnosis and timely intervention. Furthermore, the type I IFN signature, assessed through serum interferon levels, ISG transcription, and expression of interferon-stimulated proteins (*47*), may serve as a valuable marker for disease activity and severity. Notably, interferon-blocking therapies targeting IFNα protein, IFNAR, and the JAK-STAT signaling pathway are now approved for the treatment of SLE (*38*) and may also prove effective for patients with IRAK2 deficiency. Further research in animal models and preclinical trials is necessary to evaluate the long-term safety and efficacy of these targeted therapies.

In summary, we identified a loss-of-function mutation, *IRAK2-*Δ*ex2*, in three patients exhibiting immune dysregulation disorders. The *IRAK2-*Δ*ex2* mutation resulted in impairment in NF-*κ*B and MAPK signaling pathways, and upregulated type I IFN signature in both patients and mice. Our study highlighted the critical role and dual function of IRAK2 in regulating inflammatory responses and provided insights in the pathogenesis of immune dysregulation disorder due to IRAK2 deficiency.

## MATERIALS AND METHODS

### Study design

The objective of this study was to determine the correlation between IRAK2 protein deficiency and the clinical manifestation observed in three patients. Through WES and Sanger sequencing, identical breakpoints were identified in all patients each of whom was homozygous for the variation. Then, the effect of the variation on the interaction between IRAK2 and IRAK4 was confirmed through co-immunoprecipitation assay in HEK293T cells. Functional experiments were subsequently conducted using HEK293T cells, human PBMCs, and mouse BMDMs to verify the impact of IRAK2 protein deficiency on NF-*κ*B, MAPK, and type I IFN signaling pathways. The goal was to determine whether, in addition to deficiencies in the NF-*κ*B and MAPK signaling pathways, there are other abnormally activated signaling pathways present in conditions of IRAK2 protein deficiency.

### Patients

All participants, or their legal guardians, provided written informed consent. All patients enrolled in this study were evaluated following a protocol approved by the Institutional Review Boards. This study was conducted in accordance with all relevant ethical regulations.

### Cell lines, general cell culture, and treatment

The HEK293T cell line was obtained from the American Type Culture Collection (ATCC). A stable HEK293T-TLR4 cell line was established through lentiviral transduction of *TLR4-HA*, *CD14-Flag*, and *LY96*. *IRAK2* gene knockout HEK293T cells were generated using the CRISPR/Cas9 system (*21*). HEK293T cells and BMDMs were cultured in DMEM (C11995500BT, Gibco) supplemented with 10% FBS (NFBS-2500A, Noverse). PBMCs were cultured in RPMI-1640 (C11875500BT, Gibco) supplemented with 10% FBS (NFBS-2500A, Noverse).

The stable HEK293T-TLR4 cell line was stimulated with 1 µg/ml LPS (L6529, sigma) for specified durations. PBMCs were stimulated with 1 µg/ml LPS for 8 hours for single-cell RNA sequencing (scRNA-seq) and for 12 hours for CytoF and Cytometric Bead Array (BD Biosciences) at the density of 1.5e6/ml. BMDMs were stimulated with 100 ng/ml LPS, 25 ng/ml R848 (HY-13740, MCE), and 50 ng/ml FSL-1 (HY-P2036, MCE), respectively for 8 hours prior to RNA extraction.

### Mice

*Irak2^Δex2/Δex2^* mice were generated on a C57BL/6J background by GemPharmatech Co.,Ltd. (Nanjing, China) using CRISPR/Cas9 technology to delete Exon 2 of *Irak2* gene in the mouse genome. All mice were maintained in a specific pathogen-free (SPF) environment. The animals used in this study adhered to all relevant ethical regulations for animal research and were age- and sex-matched. All experimental protocols involving animals were approved by Zhejiang University’s Institutional Animal Care and Use Committee.

### Induction of BMDMs *ex vivo*

After carbon dioxide euthanasia, bone marrow cells were harvested from the femurs and tibias of wild type or *Irak2*^Δ^*^ex2/^*^Δ^*^ex2^* mice. These cells were then cultured in DMEM supplemented with 10% FBS, 1% penicillin/streptomycin (15140122, Gibco), and 10 ng/ml mouse M-CSF (315-02, PeproTech) for up to 6 days. The adherent BMDMs were subsequently utilized for further analysis.

### Whole exome sequencing

Maxwell RSC Whole Blood DNA Kit (AS1520, Promega) was employed to extract DNA from whole blood samples. One microgram of DNA was utilized for WES. The sequencing and subsequent data analysis were conducted according to previously established protocols (*48*). For analysis of single-nucleotide variants or small indels, variants were annotated utilizing ANNOVAR (2019Oct24). We removed variants presenting in the gnomAD, ClinVar, Kaviar, dbSNP, and an in-house database. Furthermore, variants were screened based on recessive or *de novo* inheritance patterns. In addition, we identified CNVs by performing the germline CNV detection pipeline of Genome Analysis Toolkit 4 (GATK4). All unaffected family members underwent genotyping through Sanger sequencing.

### Sanger sequencing

Sanger sequencing was used to confirm the CNV identified by WES.

### Antibodies and expression plasmids

The following antibodies were purchased from Cell Signaling Technology: p-p65 (Ser536) (#3033; 1:1000), p65 (#8242; 1:1000), p-p38 (Thr180/Tyr182) (#4511; 1:1000), p38 (#8690; 1:1000), HA-tag (#3724S; 1:1000), p-p105 (#4806; 1:1000), 105 (#4717; 1:1000), p-JNK (Thr183/Tyr185) (#4668; 1:1000), JNK (#9252; 1:1000). Flag-tag (MA1-91878; 1:3000) was purchased from Sigma-Aldrich. Myc-tag (60003-2-Ig; 1:3000) and HRP-conjugated GAPDH (HRP-60004; 1:3000) were purchased from Proteintech.

Human wild-type *IRAK2*, *IRAK4*, *TRAF6*, *TLR4*, *CD14*, *LY96* and the mutant *IRAK2-*Δ*ex2* were engineered by cloning the corresponding human cDNA into lenti vector which was constructed in-house with or without a tag.

### Transfection

Transient transfection of plasmids into HEK293T cells was carried out using Lipofectamine 2000 (11668019, Invitrogen), following the manufacturer’s instructions. Cells were harvested 24 to 30 hours post-transfection.

### Western blotting and co-immunoprecipitation

Cells were lysed using cold cell lysis buffer (20 mM Tris-HCl, pH 7.4, 150 mM NaCl, 0.5% NP-40) along with cOmplete protease inhibitor cocktail (Roche) and PhosSTOP (Roche). The lysate was vortexed intermittently every 10 min for a total of three times. Subsequently it was centrifuged at 12,000g for 10 min at 4 °C. The supernatant was collected for the protein concentration determination using BCA protein assay kit (23225, Thermo Fisher).

Proteins were separated on a 10% SDS-PAGE gel and then transferred onto a 0.45 µm PVDF membrane (Millipore). The membrane was then probed using specific primary and secondary antibodies. Protein bands were detected and analyzed with the Odyssey infrared imaging system (LI-COR Biosciences).

For co-immunoprecipitation, the protein concentration-adjusted cell lysates were incubated with immunomagnetic beads (B26102, Selleckchem) at 4 °C overnight. Subsequently, the beads were washed five times with lysis buffer and the complexes were analyzed by Western blotting.

### Luciferase assay

NF-*κ*B response element Firefly element reporter plasmid and Renilla luciferase expression plasmid, along with *IRAK2-WT* or *IRAK2-* Δ*ex2* expression plasmids, were co-transfected into HEK293T-*IRAK2* knock out cells. Cells were harvested 24 hours post-transfection, and luciferase activity was measured using a multimode plate reader (BioTek). Data were analyzed as fold induction, with Firefly luciferase activity normalized to Renilla luciferase activity.

### Cytokine detection

Serum or supernatant of PBMCs were analyzed using BD Cytometric Bead Array (BD Biosciences) to quantify concentrations of proinflammatory cytokines. Data analysis was conducted using the FCAP Array V3 software (BD Biosciences).

### Protein structure modeling

An illustration of the interaction between the death domains of IRAK2 and IRAK4, based on data from the Protein Data Bank (3MOP) (*3*), was prepared using UCSF ChimeraX (*19*).

### Quantitative RT-PCR

BMDMs was stimulated with either LPS (100 ng/ml), R848 (25 ng/ml), or FSL-1 (50 ng/ml) for 8 hours. Then total RNA from BMDMs was extracted utilizing the RNeasy Mini Kit (74104, Qiagen). cDNA was generated by HiScript III All-in-one RT SuperMix (R333-01, Vazyme) following manufacturer’s instruction. And qPCR was performed by using 2× Universal SYBR Green Fast qPCR Mix (RK21203, ABclonal), running on ROCHE LightCycler 480II system. Relative mRNA levels were quantified using the ΔΔ*C_t_* method.

### RNA sequencing

One microgram of RNA was used for library construction using NEBNext Ultra RNA library Prep Kit for Illumina (New England Biolabs, Ipswich, MA). Sequencing was subsequently performed on Illumina Novaseq (San Diega, CA). We used HISAT2 to map sequenced reads to the human reference genome (GRCh38). The reads numbers for each gene were counted using featureCounts. We used the R package DESeq2 to perform differential gene expression analysis.

### Single-cell RNA sequencing

10X Genomics Chromium machine was used for 8,000-10,000 single cell capture. The cDNA was subsequently amplified through PCR. Library construction and sequencing were conducted as previously described (*49*). Single-cell sequencing data were aligned with the GRCh38 human reference genome and quantified by CellRanger (version 7.1, 10X Genomics). Downstream procedure was performed by the R package Seurat (version 4.4.0). Seurat function FindAllMarkers and FindConservedMarkers (both with default parameter) were used to identify differentially expressed genes between clusters for cell annotation. Human myeloid markers were collected from the previous study (*24*).

### CyTOF

PBMCs from two patients and three unaffected controls were either treated with 1 µg/ml LPS or balanced overnight prior to analysis. For mass cytometry analysis, purified antibodies were purchased from Biolegend, Thermo Fisher, Bio-Rad and R&D systems using the clones detailed in table S4. Sample preparation, flow cytometer setup and operation procedure.

FCS file processing of CyTOF and cell clustering analysis were performed according to the previous pipeline (*49*).

### The algorithm of IFN score

Firstly, 28 genes related to interferon response were selected from the previous study (*23*). They were: *CXCL10*, *DDX60*, *EPSTI1*, *GBP1*, *HERC5*, *HERC6*, *IFI27*, *IFI44*, *IFI44L*, *IFI6*, *IFIT1*, *IFIT2*, *IFIT3*, *IFIT5*, *ISG15*, *LAMP3*, *LY6E*, *MX1*, *OAS1*, *OAS2*, *OAS3*, *OASL*, *RSAD2*, *RTP4*, *SIGLEC1*, *SOCS1*, *SPATS2L*, and *USP18*. Secondly, we calculated the z-scores of each 28 genes with DESeq2 normalized reads counts using the mean and standard deviation of controls. Subsequently, for each sample, we summed the 28 z-scores as their IFN response score.

### Calculation of signature score for scRNA-seq

To evaluate the immune response among individuals for the given cell type, we used the R package AUCcell (*26*) to calculate to signature score for NF-*κ*B and type I IFN. AUCcell is ranked-based, which is independent of gene expression units and the normalized method. The NF-*κ*B gene set was obtained from the resource (https://www.bu.edu/nf-kb/gene-resources/target-genes/), including cytokines, chemokines, immune receptors, etc. Type I IFN gene set was collected from previous report (*23*).

### Flow cytometry analysis of phosphorylation

For detection of phosphorylated protein, isolated PBMCs were permeabilized using Perm Buffer III (558050, BD Biosciences). Cells were then stained with antibodies against CD16 (563692, BD Bioscience), and p-STAT1 (pY701) (612564, BD Bioscience). Data were analyzed using FlowJo.

### Statistical analysis

GraphPad Prism version 9.5.0 for Mac OS X, GraphPad Software, Boston, Massachusetts USA, www.graphpad.com, was used for statistical analysis. For variables following a normal distribution, comparisons between two groups were performed using the Student’ s *t*-test (unpaired and two-tailed). P values < 0.05 were considered statistically significant. In figures, asterisks denote statistical significance (\**P* < 0.05; \*\**P* < 0.01; \*\*\**P* < 0.001). Statistical analysis of single-cell RNA sequencing, CytoF, and RNA sequencing was performed using R Software (R v.4.4.0).

Molecular graphics and analyses performed with UCSF ChimeraX, developed by the Resource for Biocomputing, Visualization, and Informatics at the University of California, San Francisco, with support from National Institutes of Health R01-GM129325 and the Office of Cyber Infrastructure and Computational Biology, National Institute of Allergy and Infectious Diseases.

The flow cytometry results were analyzed using FlowJo™ v10.8 Software (BD Life Sciences).

## List of Supplementary Materials

Figs. S1 to S3

Tables S1 to S4

## Acknowledgments

We thank the patients, their families and the unaffected controls for their support during this research study.

## Funding

The work was supported by the National Natural Science Foundation of China (82225022, 32141004, 82394424, 32321002, 8240062882, 82170739 and 82402088), the Hundred-Talent Program of Zhejiang University, Leading Innovative and Entrepreneur Team Introduction Program of Zhejiang (Grant No. 2021R01012), the National key research and development project of China (Grant No. 2021YFC2501302), the Open Project of Jiangsu Provincial Science and Technology Resources (Clinical Resources) Coordination Service Platform (Grant No. TC2023B004), the Postdoctoral Fellowship Program of CPSF (Grant No. GZB20230635), and the China Postdoctoral Science Foundation (Grant No. 2024T170779, 2024M752825).

## Author contributions

Conceptualization, Funding acquisition, Project administration & Supervision: Zhihong Liu, Qing Zhou, Xiaomin Yu,

Functional experiment: Yudie Fei, Lin Liu, Shihao Wang, Yusha Wang, Xiangwei Sun,

Data analysis: Shuangyue Ma, Xiang Chen, Xu Han, Jiahui Zhang,

Patient clinical information: Meiping Lu, Jing Xue, Ying Jin, Li Guo,

Writing – original draft: Yudie Fei

Writing – review & editing: Lin Liu, Shuangyue Ma, Shihao Wang, Yusha Wang, Xu Han, Qing Zhou

## Competing interests

Authors declare that they have no competing interests.

## Data and materials availability

All data are available in the main text or the supplementary materials.

## Notes

### Competing Interest Statement

The authors have declared no competing interest.

### Funding Statement

The work was funded by the National Natural Science Foundation of China (82225022, 32141004, 82394424, 32321002, 8240062882, 82170739 and 82402088), the Hundred-Talent Program of Zhejiang University, Leading Innovative and Entrepreneur Team Introduction Program of Zhejiang (Grant No. 2021R01012), the National key research and development project of China (Grant No. 2021YFC2501302), the Open Project of Jiangsu Provincial Science and Technology Resources (Clinical Resources) Coordination Service Platform (Grant No. TC2023B004), the Postdoctoral Fellowship Program of CPSF (Grant No. GZB20230635), and the China Postdoctoral Science Foundation (Grant No. 2024T170779, 2024M752825).

### Author Declarations

Institutional Review Boards of Zhejiang university and National Clinical Research Center of Kidney Diseases Jinling Hospital gave ethical approval for this work.

