## supplement Figs. S1 to S3 Tables S1 to S4 for "IRAK2 deficiency causes a new immune dysregulation disorder"

**List of Supplementary Materials**

Figs. S1 to S3

Tables S1 to S4

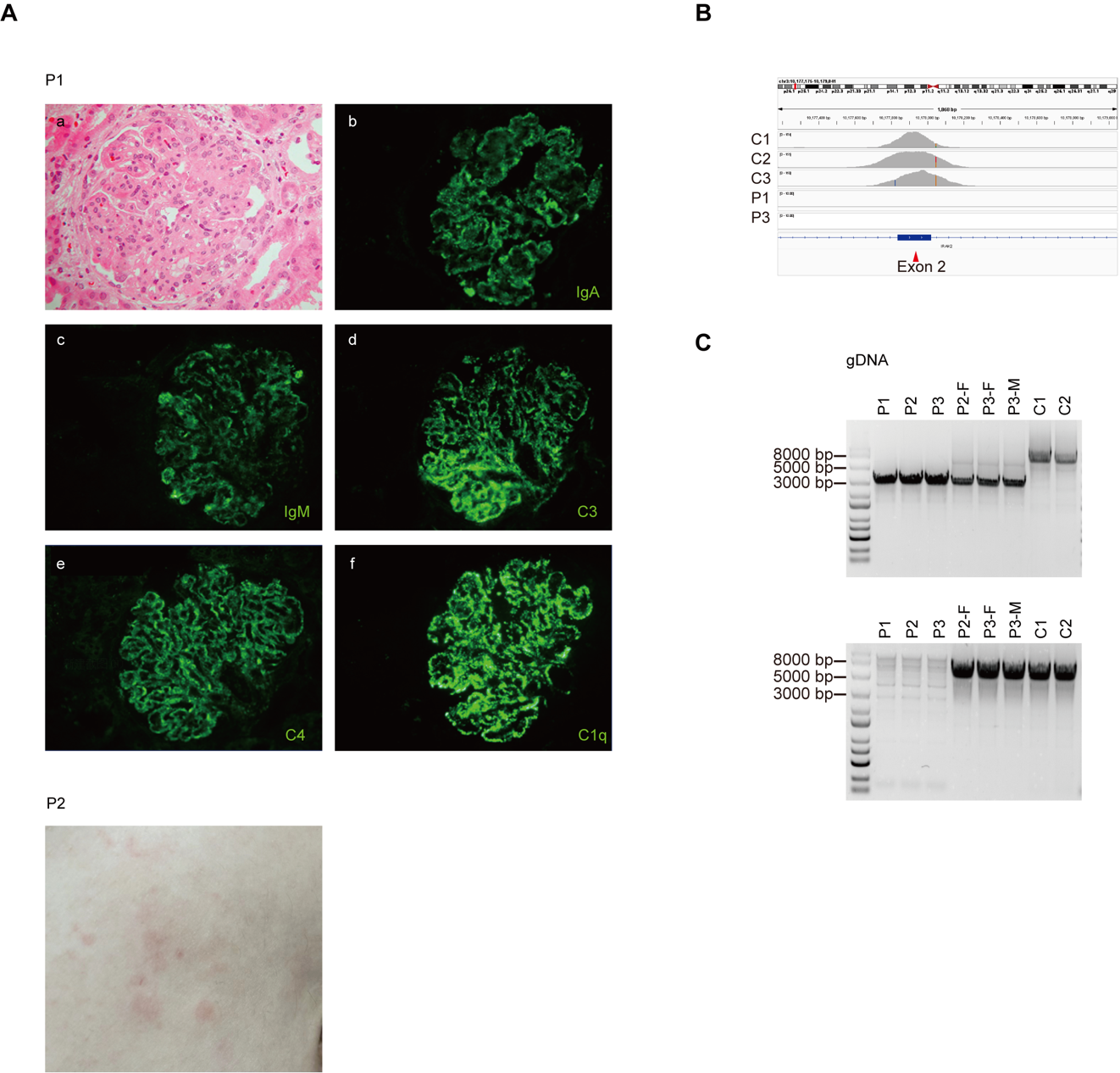

**Figure S1. Clinical manifestations of P1 and P2 and confirmation of the *IRAK2* variant.** (A) Renal pathological examination of P1 and skin rashes of P2. Light microscopy: (a) Neutrophil infiltration and mesangial as well as endocapillary proliferation in the glomerulus (HE, ×400). Immunofluorescence: (b-f) Diffusely distributed deposits of IgA, IgM, C3, C4, C1q in a granular pattern within the vascular loops (IF, ×400). (B) Exon sequencing reads covering the *IRAK2-∆ex2* variant in P1, P3 and three unaffected controls, as displayed by the Integrative Genomics Viewer. (C) Agarose gel electrophoresis results of PCR amplification products generated using primers flanking two breaking points (above) and with one primer located inside the breaking points (below), using gDNA as templates.

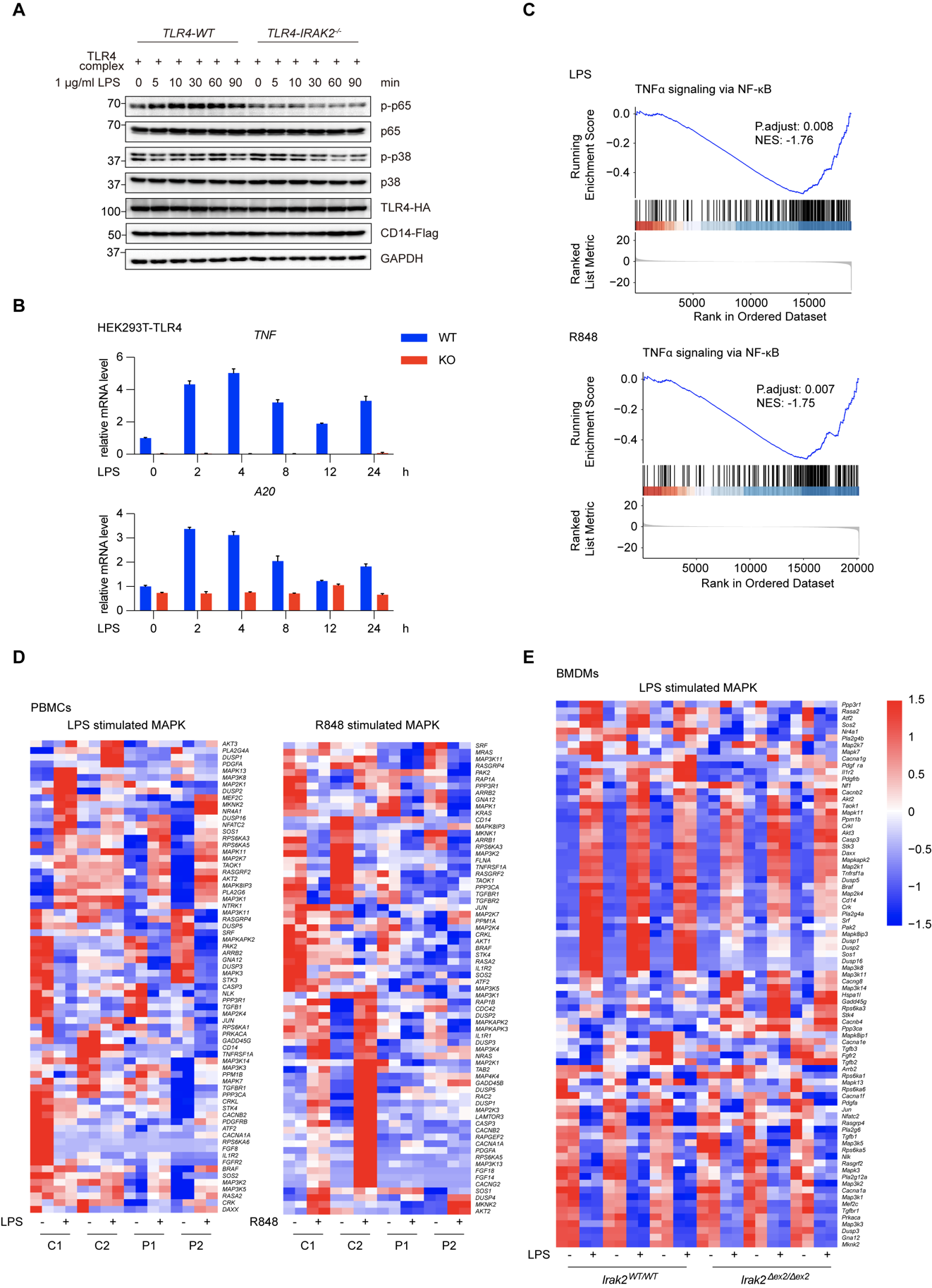

**Figure S2. Deficiency in the TLR signaling pathways via Myddosome.** (A) Immunoblot analysis demonstrating deficiencies in NF-𝜅B and MAPK signaling in HEK293T-*IRAK2^-/-^* cells compared with wild-type. (B) Quantitative PCR (qPCR) analysis of the expression of the genes related to NF-𝜅B signaling in HEK293T-*IRAK2^-/-^* cells and wild-type HEK293T, treated with 100 ng/ml LPS for the indicated times or left untreated. (C) Gene set enrichment analysis plots of differentially expressed genes from RNA sequencing data in PBMC treated with 1 μg/ml LPS (above) and 1 μg/ml R848 (below) respectively for 12 hours. (D) RNA sequencing analysis of MAPK pathway in PBMCs treated with 1 μg/ml LPS, 1 μg/ml R848, respectively, or left untreated for 12 hours. (E) RNA sequencing analysis of MAPK pathway in BMDMs treated with 100 ng/ml LPS or left untreated for 8 hours.

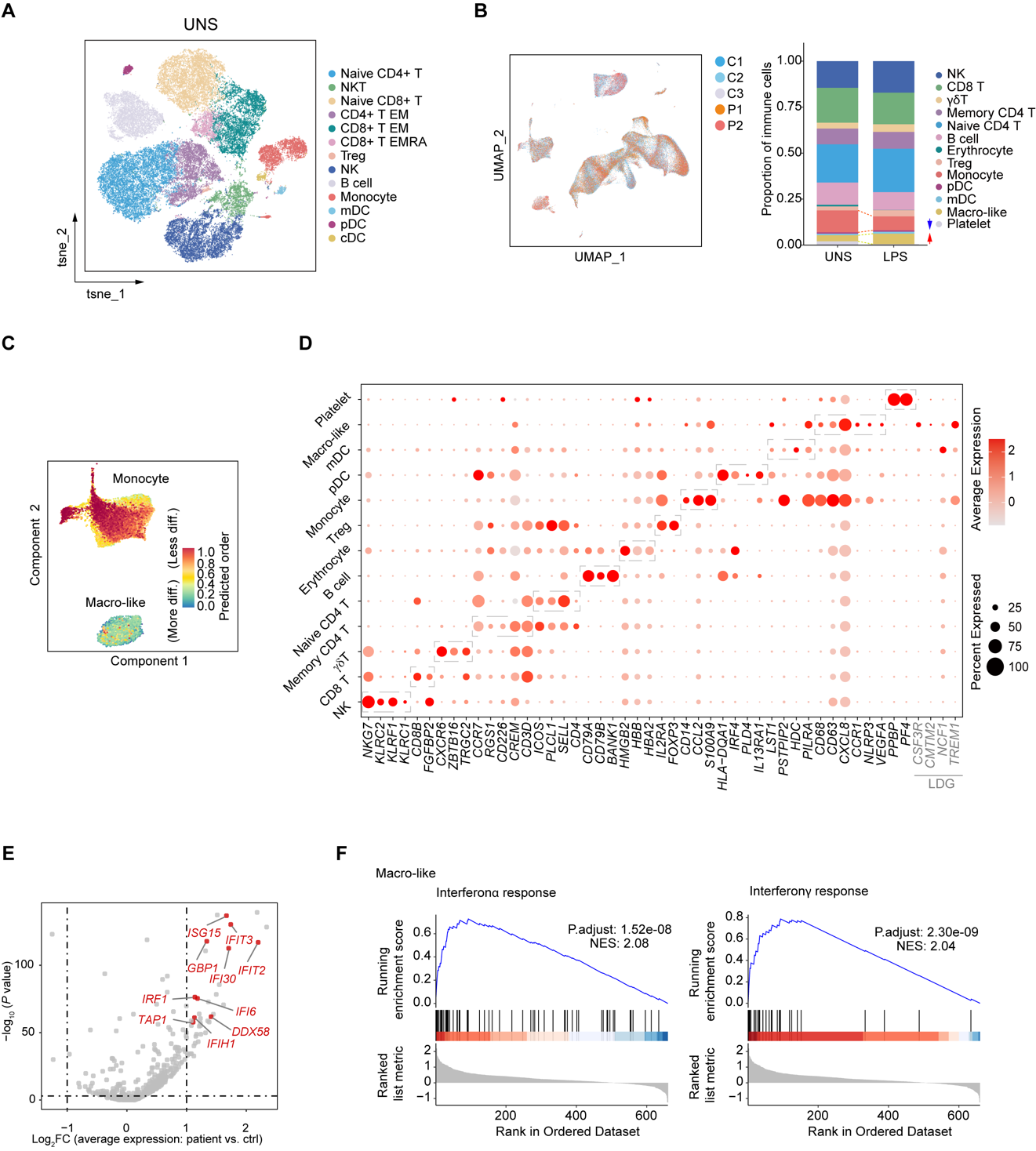

**Figure S3. Results of CyTOF and scRNA-seq using PBMCs from P1, P2, and unaffected controls.** (A) The t-distributed stochastic neighbor embedding (t-SNE) representation illustrates the immunophenotyping of 5*10,000 CD45^+^ CD66b^-^ live PBMCs balanced overnight from two patients and three unaffected controls, as analyzed by CyTOF. (B) Uniform Manifold Approximation and Projection (UMAP) visualization displays cell distribution corresponding to P1, P2, and three unaffected controls (group.by = “sample”). Cell proportions are displayed as a stacked bar plot. (C) Differentiation states of monocytes and ‘Macro-like’ cell subset assessed by CytoTRACE algorithm (*25*). (D) The bubble graph shows the expression of marker genes for annotating cell components of PBMCs by scRNA-seq. (E) Volcano plot depicting differential gene expression between ‘Macro-like’ cell subset from patients and unaffected controls upon LPS stimulation. (F) Gene set enrichment analysis plots of differentially expressed genes identified between patients and unaffected controls in ‘Macro-like’ cell subset based on scRNA-seq data (LPS treated).

**Table S1 Laboratory features of the patients.**

|  | F1. P1 | F1. P2 | F2. P3 | Reference value |
| --- | --- | --- | --- | --- |
| Diagnosed age (y) | 21-25 |  | 11-15 | \ |
| Current age (y) | 46-50 | 21-25 | 21-25 | \ |
| Sex | Female | Female | Male | \ |
| White blood cell (10^9/L) | 5.9 | 5.57 | 10.940↑ | 3.5-9.5 |
| Hemoglobin (g/L) | 95↓ | 150 | 115 | 115-150 |
| Platelet (10^9/L) | 146 | 190 | 431↑ | 100-300 |
| Serum creatinine (μmol/L) | 147 ↑ | 62 |  | 45-110 |
| Blood urea nitrogen (mmol/L) | 9 ↑ | 4.49 |  | 2.9-7.5 |
| ANA | 1:320↑ | 1:160 ↑ | <1:40 | <1:40 |
| Anti-dsDNA | <1:10 | <1:10 | <1:10 | <1:10 |
| C3 (g/L) | 0.526 ↓ | 0.8 | 1.23 | 0.8-1.8 |
| C4 (g/L) | 0.117 | 0.108 | 0.25 | 0.1-0.4 |
| Anti-β2 glycoprotein I (RU/ml) |  | 157.78 ↑ |  | <20 |
| ACL IgG (U/ml) | Negtive | 88.86 ↑ |  | <15 |
| ACL IgM (U/ml) | Negtive | 46.29 ↑ |  | <15 |
| ACL IgA (U/ml) | Negtive | Negtive |  | <15 |
| CRP (mg/L) |  |  | 104↑ | <10 |
| ESR (mm/h)  IgG (g/L)  IgM (g/L)  IgA (g/L) | 6.25↓  0.256↓  0.041↓ |  | 8 | 0-20 |
| IgG (g/L) | 6.25↓ |  |  | 7-16 |
| IgM (g/L) | 0.256↓ |  |  | 0.4-2.3 |
| IgA (g/L) | 0.041↓ |  |  | 0.7-4.0 |

**Table S2 Laboratory features of the patient P2.**

| F1. P2 | 2022 | 2023_1 | 2023_2 | 2024 | Reference value |
| --- | --- | --- | --- | --- | --- |
| White blood cell (10^9/L) | 5.57 | 6.06 |  | 7.61 | 3.5-9.5 |
| Hemoglobin (g/L) | 150 | 155↑ |  | 145 | 115-150 |
| Platelet (10^9/L) | 190 | 204 |  | 199 | 100-300 |
| Serum creatinine (μmol/L) | 62 | 56.6 | 56.5 | 63.6 | 45-110 |
| Blood urea nitrogen (mmol/L) | 4.49 | 2.64↓ | 2.96 | 4.14 | 2.9-7.5 |
| ANA | 1:160 ↑ | 1:320↑ | 1:640 ↑ | 1:1280 ↑ | <1:40 |
| Anti-dsDNA | <1:10 | <1:10 | <1:10 | <1:10 | <1:10 |
| C3 (g/L) | 0.800 | 0.788↓ | 0.880 | 0.885 | 0.8-1.8 |
| C4 (g/L) | 0.108 | 0.112 | 0.125 | 0.123 | 0.1-0.4 |
| Anti-β2 glycoprotein I (RU/ml) | 157.78 ↑ | 266↑ | >300↑ | >300 ↑ | <20 |
| ACA (RU/ml) | Positive | >300↑ | >300↑ | >300↑ | <12 |
| ACL IgG (U/ml) | 88.86↑ |  |  | >120 ↑ | <15 |
| ACL IgM (U/ml) | 46.29↑ |  |  | 24.5 ↑ | <15 |
| ACL IgA (U/ml) | Negative |  |  | 8.8 | <15 |
| CRP (mg/L) | <10 |  |  | <10 | <10 |

**Table S3. Flow cytometry results for P1 and P2.**

| Cell type | Surface marker | Denominator | Test value % (P1) | Test value % (P2) | Reference % |
| --- | --- | --- | --- | --- | --- |
| Granulocytes | CD45+SS (high) | White blood cells | 61.68 | 50.07 | 37.20-80.50 |
| Monocytes | CD14+ | White blood cells | 6.67 | 7.56 | 3.00-10.90 |
| Classical monocytes | CD14highCD16- | Monocytes | 88.94 | 71.16 | 68.44-93.40 |
| Intermediate monocytes | CD14highCD16+ | Monocytes | 6.77 | 12.26 | 2.60-15.80 |
| Proinflammatory monocytes | CD14+CD16high | Monocytes | 0.98↓ | 11.45 | 2.20-16.70 |
| Dendritic cells (DC) | Lin-HLA-DR+ | White blood cells | 0.22 | 0.24 | 0.20-1.90 |
| Plasmacytoid dendritic cells (pDC) | Lin-HLADR+CD11c-CD123+ | White blood cells | 0.03 | 0.09 | 0.00-0.40 |
| Myeloid dendritic cells (mDC) | Lin-HLA-DR+CD11c+ | White blood cells | 0.15 | 0.13 | 0.10-1.70 |
| CD16 positive myeloid dendritic cells | HLADR+CD11c+CD16+ | mDC | 40.38 | 24.62↓ | 33.90-98.20 |
| Myeloid dendritic cells subtype 1 (MDC1) | Lin-HLADR+CD11c+CD16-CD1c+Clec9A- | mDC | 44.81 | 58.96 | 1.70-61.60 |
| Myeloid dendritic cells subtype 2 (MDC2) | Lin-HLADR+CD11c+CD16-CD1c-Clec9A+ | mDC | 1.54 | 1.68 | 0.10-4.50 |
| Lymphocytes | CD45+SS (low) FS (low) | White blood cells | 31.21 | 40.26 | 11.40-57.00 |
| Natural killer cells (NK cells) | CD3-CD56+ | Lymphocytes | 20.11 | 7.72 | 3.30-32.90 |
| Mature natural killer cells | CD3-CD56dim | NK cells | 96.13 | 94.67 | 71.90-99.50 |
| Immature natural killer cells | CD3-CD56high | NK cells | 3.87 | 5.33 | 0.50-28.10 |
| NKT cells | CD3+CD56+ | Lymphocytes | 5.36 | 2.97↓ | 3.00-8.00 |
| Total T cells | CD3+ | Lymphocytes | 76.30 | 68.45 | 53.70-82.80 |
| Helper T cells | CD3+CD4+ | T cells | 33.61↓ | 34.62↓ | 46.20-78.00 |
| Activated CD4+ T cells | CD3+CD4+HLA-DR+ | CD4+ T cells | 14.24 | 5.31 | 3.60-31.40 |
| CD4+ naïve T cells | CD3+CD4+CCR7+CD45RA+ | CD4+ T cells | 19.81 | 43.62 | 7.20-68.90 |
| CD4+ central memory T cells | CD3+CD4+CCR7+CD45RA- | CD4+ T cells | 61.88 | 35.48 | 15.00-64.30 |
| CD4+ effector memory T cells | CD3+CD4+CCR7-CD45RA- | CD4+ T cells | 17.96 | 20.78 | 3.13-24.83 |
| CD4+ terminally differentiated effector memory T cells | CD3+CD4+CCR7-CD45RA+ | CD4+ T cells | 0.35 | 0.12↓ | 0.16-14.41 |
| Regulatory T cells (Treg) | CD3+CD4+CD25highFoxP3+ | CD4+ T cells | 3.24↓ | 5.08↓ | 5.10-12.70 |
| Naïve regulatory T cells | CD3+CD4+CD25highFoxP3+CD45RA+ | Treg cells | 0.81↓ | 25.11 | 3.50-77.30 |
| Memory regulatory T cells | CD3+CD4+CD25highFoxP3+CD45RA- | Treg cells | 99.19↑ | 74.89 | 22.70-96.50 |
| Natural regulatory T cells (nTreg) | CD3+CD4+CD25highFoxP3+Hellios+CD39+ | CD4+ T cells | 1.78 | 1.46 | \ |
| Induced regulatory T cells | CD3+CD4+CD25highFoxP3+Hellios- | CD4+ T cells | 0.10 | 1.25 | \ |
| Cytotoxic T cells | CD3+CD8+ | T cells | 62.30↑ | 60.53↑ | 14.80-48.40 |
| Activated CD8+ T cells | CD3+CD8+HLA-DR+ | CD8+ T cells | 35.78 | 12.84 | 6.10-63.50 |
| CD8+ naïve T cells | CD3+CD8+CCR7+CD45RA+ | CD8+ T cells | 46.07 | 48.81 | 2.60-72.40 |
| CD8+ central memory T cells | CD3+CD8+CCR7+CD45RA- | CD8+ T cells | 10.03 | 3.99 | 2.70-36.20 |
| CD8+ effector memory T cells | CD3+CD8+CCR7-CD45RA- | CD8+ T cells | 17.38 | 38.02 | 4.70-60.10 |
| CD8+ terminally differentiated effector memory T cells | CD3+CD8+CCR7-CD45RA+ | CD8+ T cells | 26.52 | 9.18 | 1.60-62.00 |
| Double negative T cells | CD3+CD4-CD8- | T cells | 2.79 | 3.74 | \ |
| Double positive T cells | CD3+CD4+CD8+ | T cells | 1.25 | 0.76 | \ |
| γδT cells | CD3+TCRγδ+ | T cells | 2.52 | 4.56 | 0.40-20.70 |
| TCRγδ1 T cells | CD3+TCRγδ+ TCRγδ1+ | γδT cells | 55.21↑ | 41.31↑ | 1.00-30.00 |
| TCRγδ2 T cells | CD3+TCRγδ+ TCRγδ2+ | γδT cells | 37.92↓ | 36.69↓ | 50.00-90.00 |
| Total B cells | CD19+CD3- | Lymphocytes | 0.72↓ | 20.76 | 3.80-21.50 |
| Transitional B cells | CD19+Cd27-CD38high IgM+CD24+ | B cells | 0.00↓ | 0.59 | 0.10-6.30 |
| Naïve B cells | CD19+CD27-IgD+ | B cells | 17.68↓ | 83.51↑ | 17.90-75.10 |
| Marginal zone B cells | CD19+CD27+IgD+ | B cells | 12.12 | 5.50 | 3.80-35.60 |
| Memory B cells | CD19+CD27+CD38dim | B cells | 60.10↑ | 9.38↓ | 11.00-46.60 |
| Classical immunoglobulin class-converted B cells | CD19+CD27+CD38dimIgD-IgM- | B cells | 42.93↑ | 3.30↓ | 4.60-35.50 |
| Non-immunoglobulin class-converting B cells | CD19+CD27+CD38dimIgM+ | B cells | 17.17 | 5.81 | 1.90-23.70 |
| Plasmablasts | CD19+CD27highCD38highIgD-IgM- | B cells | 0.92 | 0.09↓ | 0.30-7.80 |
| CD21 low B cells | CD19+CD38lowCD21low | B cells | 9.60 | 3.28 | 1.90-19.30 |

The tests were done by KingMed Diagnostics.

**Table S4. Antibodies used for mass cytometry analysis.**

| Mass and Tag | Marker | Clone | Staining |
| --- | --- | --- | --- |
| 89Y | CD45 | HI30 | Surface |
| 115In | CD3 | UCHT1 | Surface |
| 141Pr | CD62L | DREG-56 | Surface |
| 142Nd | TCR γ/δ | 5A6.E9 | Surface |
| 143Nd | CD1c | L161 | Surface |
| 144Nd | CXCL10(IP-10) | J034D6 | Intra |
| 145Nd | CD27 | O323 | Surface |
| 146Nd | TNF-α | MAb11 | intra |
| 147Sm | CD20 | 2H7 | Surface |
| 148Nd | CD19 | HIB19 | Surface |
| 149Sm | CD25(IL-2Rα) | 24212 | Surface |
| 150Nd | CD11c | Bu15 | Surface |
| 151Eu | CD38 | HIT2 | Surface |
| 152Sm | CD45RA | HI100 | Surface |
| 153Eu | CD123(IL-3Rα) | 6H6 | Surface |
| 154Sm | CD197(CCR7) | G043H7 | Surface |
| 155Gd | CD15(SSEA-1) | W6D3 | Surface |
| 156Gd | IL-6 | MQ2-13A5 | Intra |
| 157Gd | IL-4 | MP4-25D2 | intra |
| 158Gd | STAT3 Phospho(Tyr705) | 13A3-1 | Intra |
| 159Tb | CD56 | NCAM16.2 | Surface |
| 160Gd | IL-8 | E8N1 | Intra |
| 161Dy | IL-17A | BL168 | intra |
| 162Dy | IL-1 β(IL-1F2) | 8516 | Intra |
| 163Dy | IL-2 | MQ1-17H12 | Intra |
| 164Dy | CD141(Thrombomodulin) | M80 | Surface |
| 165Ho | IFN-γ | B27 | Intra |
| 166Er | IL-23(p19) | HLT2736 | intra |
| 167Er | STAT1 Phospho(Ser727) | A15158B | Intra |
| 168Er | GM-CSF | BVD2-21C11 | Intra |
| 169Tm | CD33 | WM53 | Surface |
| 170Er | CD66b | G10F5 | Surface |
| 171Yb | CD127(IL-7Rα) | A019D5 | Surface |
| 172Yb | HLA-DR | L243 | Surface |
| 173Yb | IL-10 | JES3-9D7 | Intra |
| 174Yb | CD14 | M5E2 | Surface |
| 175Lu | CD16 | 3G8 | Surface |
| 176Yb | Interferon α(IFN-α) | 144BS | Intra |
| 197Au | CD4 | RPA-T4 | Surface |
| 198Pt | CD8a | RPA-T8 | Surface |
| 209Bi | CD11b | M1/70 | Surface |
